# Parenthood and longitudinal changes in alcohol use – findings from two follow-up studies

**DOI:** 10.1101/2025.09.10.25335507

**Authors:** Jenna Grundström, Olli Kiviruusu, Hanna Konttinen, Sari Aaltonen, Antti Latvala, Noora Berg

## Abstract

The transition to parenthood is a significant life event that can influence various aspects of individuals’ lives, including alcohol consumption patterns in long-term. This study examines alcohol use changes in relation to parenthood in two Finnish cohorts.

Data were derived from two follow-up studies: the TAM cohort (ages 16, 22, 32, 42, and 52) and the FinnTwin16 cohort (16, 18, 25 and 35). We used generalized linear mixed model to examine the association of parenthood years and alcohol use from adolescence to mid-adulthood. Frequent alcohol use (FAU) and heavy episodic drinking (HED) were examined separately.

FAU increased with age in both cohorts, while HED peaked in late adolescence among women and in young adulthood among men. Prior to parenthood, alcohol use was generally lower in the more distant years and increased as transition approached in FinnTwin16. HED declined in the early years following the transition to parenthood, while FAU remained stable. In both women and men, long-term increases in alcohol use were observed around seven years after becoming a parent. Differences between parents and nonparents were limited.

These findings suggest that parenthood is associated especially with short-term reductions in HED, but that overall alcohol use tends to increase over time.

## 1 Introduction

Becoming a parent marks a profound shift in roles, responsibilities, and daily life, encompassing both joy and stress for new parents (Nomaguchi & Milkie, 2003; Asselmann et al., 2020). Amidst these changes, individuals navigate the complexities of parenthood, wherein their alcohol consumption habits may also undergo notable changes. While existing studies have predominantly focused on pregnancy or immediate post-transition changes in alcohol use (Liu et al., 2015; Mellingen et al., 2013; Tung et al., 2020), these changes can also be long-term, given the responsibilities and caregiving demands or changes in priorities that continue beyond the initial transition. For some individuals, the demands of parenthood may increase alcohol consumption as they may seek temporary relief from the pressures of caregiving (Caldeira & Woodin, 2012). Conversely, the social expectations and time constraints imposed by parenthood may limit opportunities to engage in such behaviors and change leisure activities, such as social gatherings with alcohol (Laborde & Mair, 2012; Paradis et al., 2011). As children grow, the responsibilities might change and parents may engage in more frequent alcohol use (Leggat et al., 2021). This study aims to examine the life course trajectories of alcohol use in the context of parenthood exploring how alcohol use develops before parenthood and how parents may change their alcohol use (frequent alcohol use and heavy episodic drinking) over time starting from adolescence and lasting up to mid-adulthood years. Specifically, we used two Finnish cohort studies to examine alcohol use trajectories before and after first parenthood, allowing us to assess cohort effects and improve the generalizability of our finding. Additionally, we examined the differences in alcohol use trajectories between parents and non-parents from adolescence to mid-adulthood.

### 1.1 Parenthood and the trajectory of alcohol use

From the life course perspective, becoming a parent presents an important life transition that can significantly alter different life trajectories. Becoming a parent is not seen as a singular event, but as a continuous process that influences one’s behavior and lifestyle choices, such as alcohol consumption. Research has shown that parents tend to consume less alcohol compared to non-parents, highlighting the transformative association of parenthood on lifestyle choices (Evans-Polce et al., 2020; Fergusson et al., 2012). However, the patterns of alcohol use among parents can still fluctuate significantly throughout the life course.

Previous studies have predominantly focused on the initial changes in alcohol use during the pregnancy (Ethen et al., 2009; Kesmodel & Urbute, 2019; Mellingen et al., 2013; Nilsen et al., 2008) or the immediate years following childbirth (Laborde & Mair, 2012; Richman et al., 1995; Tung et al., 2020), capturing the immediate responses to new responsibilities of parenting. Most of these studies examined only women, and commonly report a reduction in alcohol use during pregnancy (Leggat et al., 2021; Mellingen et al., 2013), possibly due to well-known dangers of alcohol use to the fetus. In the postnatal period, however, the studies have yielded some mixed results. While some studies indicate a continued decrease in alcohol use five years after becoming a parent (Laborde & Mair, 2012; ref.), other studies have noted a significant increase in alcohol consumption during this phase, especially among men (Axinn et al., 2022; Leggat et al., 2021).

Studies that have found a decreasing trajectory of alcohol use after the transition, have also suggested that this association might be relatively short-lived (Borschmann et al., 2019). An immediate reduction in alcohol use after becoming a parent might return to pre-parenting levels years after this (Bailey et al., 2008; Borschmann et al., 2019), therefore concluding that the influence of becoming a parent does not have a lasting association on alcohol consumption patterns within parents. However, the long-term changes in alcohol consumption or the trajectory of alcohol use in relation to parenthood – both before and after becoming a parent – are less studied. As children grow and family dynamics evolve, parents may face different stressors and social situations, potentially influencing their drinking behaviors over the long-term. Previous research has studied the relationship between the parenting stage – defined by the age of child or children – and alcohol consumption. These studies have suggested that parents with younger children tend to drink less compared to those with older children or those without children, especially among women (Borschmann et al., 2019; Wolfe, 2009). These findings propose that while becoming a parent often initially leads to decrease in alcohol use, this association may diminish over time, with the risk of alcohol abuse and binge drinking increasing as children age, eventually reverting to levels seen prior to becoming a parent. However, a comprehensive examination of the longitudinal trajectories of alcohol use from adolescence through mid-adulthood remains scarce.

When comparing the patterns of alcohol use of parents and nonparents, parenthood has often been associated with reduced alcohol consumption, particularly among women (Christie-Mizell & Peralta, 2009). This observed decrease in alcohol intake among parents may be attributed to various factors inherent to the parenthood, like heightened sense of responsibility and the need to prioritize childcare duties, leading parents to adopt healthier lifestyle choices that include reduced alcohol intake. For example, Matusiewicz et al. (2016) studied the changes in mothers’ alcohol use over three years during the transition to parenthood and compared them with those who did not became mothers. Study found similar levels of alcohol use at baseline but significant decreases in alcohol use after the transition among mothers, whereas those who did not become mothers, reported increases in the frequency of alcohol use.

Patterns of alcohol use can be examined across multiple dimensions, including usage frequency and the occurrence of heavy episodic drinking. Existing studies have revealed that different measures of alcohol use can exhibit different trends (Christie-Mizell & Peralta, 2009; Evans-Polce et al., 2020), although similar trends have also been found (Leggat et al., 2021; Little et al., 2009). Frequency of alcohol use refers to how often individuals consume alcohol over a given period. Research indicates that becoming a parent often leads to a reduction in drinking frequency, due to the increased responsibilities and time constraints associated with childcare (Staff et al., 2010). On the other hand, heavy episodic drinking (HED) is defined as consuming a large quantity of alcohol on one occasion. After becoming a parent, heavy episodic drinking may become less frequent as parents might not have similar opportunities for heavy episodic drinking, such as drinking in a bar or pubs (Paradis, 2011). Leggat et al. (2021) found a distinction between these two dimensions of alcohol use. Their study reported an increase in the usual quantity of alcohol consumed after childbirth among mothers and fathers, but not in the frequency of drinking occasions. This suggests that while parents may drink less often, the amount they consume when they do drink can be significant. This pattern highlights the need to differentiate between various types of alcohol consumption behaviors when studying the impact of parenthood.

#### 1.1.1 Gender differences

Gender differences in alcohol use have been widely documented, with numerous studies indicating that men and women exhibit distinct patterns of alcohol consumption (e.g., White, 2020). Research consistently shows that men generally consume more alcohol than women and are more likely to engage in heavy episodic drinking (White, 2020; Wilsnack et al., 2009). The association of the transition to parenthood on alcohol use is also gendered (Nomacuchi & Milkie, 2020), with women reacting more strongly to the transition to parenthood than men (Asselmann et al., 2022; Metzger & Gracia, 2022; Parfitt & Ayers, 2014). These differences may reflect the complex interplay of biological factors, societal norms, and gender roles. For example, the transition to parenthood has been quite consistently associated with less alcohol consumption among women (Dash et al., 2020) – especially during pregnancy (Bachman et al. 2002; Leggat et al. 2021). Physiological and biological factors are likely to play a central role in this due to significant risks related to alcohol use for the child both during and after pregnancy (e.g., during breastfeeding). Conversely, for men the transition to parenthood and fatherhood itself may be shaped less by biological factors and more by social expectations, such as changes in identity and social roles, and financial responsibilities. While the transition to fatherhood often entails a reevaluation of personal habits and lifestyle, the patterns observed in men’s alcohol use are less straightforward. Some studies suggest a general decline in frequent and heavy drinking among new fathers, though this reduction is typically less pronounced than that observed in women (Condon et al., 2004). However, other studies have indicated an increase in alcohol use among men during the postpartum period (Axinn et al., 2022; Leggat et al., 2021). Additionally, some studies have also found that fathers’ alcohol consumption does not seem to affected by parenthood or the transition to parenthood (Dash et al., 2020). It should be noted, however, that most studies have primarily focused on only maternal alcohol use, with less research dedicated to paternal alcohol use.

### 1.2 Present study

The present study examines the relationship between parenthood and alcohol use, focusing on patterns before and after individuals become parents. The transition to parenthood is a significant life event with potential implications for alcohol use both short- and long-term throughout the life course. However, most studies on alcohol use before and after becoming a parent have focused on the perinatal period in women, leaving a limited understanding of how these patterns change over the longer term. Using data from two Finnish cohort-studies from young adulthood to adulthood/mid-adulthood, this study aims to fill this gap by examining alcohol use trajectories years before and after parenthood, encompassing both women and men. Additionally, this study examines the differences in alcohol use between parents and non-parents across the life course. Such comparisons between parents and non-parents can provide valuable contextual information for understanding patterns of alcohol use. Women and men are studied separately due to gender differences found in previous studies on alcohol use and parenthood. By examining both frequent alcohol use and heavy episodic drinking, we aim to provide a comprehensive overview of how parenthood is associated with changes in drinking behaviors over the life course.

In summary, our research questions are:

1. How do patterns of alcohol use, specifically the frequent alcohol use and heavy episodic drinking, change before and after the transition to parenthood among women and men during the life course?
2. How do parents and non-parents differ in alcohol use among women and men throughout the life course?

## 2 Methods

### 2.1 The two cohort studies

In the present study, two longitudinal follow-up studies from Finland were used to examine trajectories of alcohol use before and after the transition to parenthood.

#### 2.1.1 TAM

The original study population in the Finnish cohort study ‘Stress, development and mental health (TAM)’ included all Finnish-speaking ninth-grade students in Tampere in 1983. A questionnaire was completed by 2,194 pupils (97% of the target population, mean age 15.9) during a school day. The participants were then followed up in 1989 (N = 1656; 76%), 1999 (N = 1471; 67%), 2009 (N = 1334; 61%), and 2019 (N = 1160; 53%), when the participants were approximately 22, 32, 42, and 52 years old (Berg et al., 2021). In each study wave, the original study population from the 1983 baseline was contacted. In the present study, only those who participated in the 2009 follow-up survey at age 42, were included (N = 1334). The 2009 follow-up survey was the last study wave when children’s birth years were inquired in the questionnaire and enabled us to calculate the time of parenthood.

The TAM study has been approved by the Ethics Committee of Tampere University Hospital and the Institutional Review Board of the Finnish Institute for Health and Welfare (THL).

#### 2.1.2 FinnTwin16

The ‘FinnTwin16’(FT16) is a population-based study of twins born in 1974-1979. FinnTwin16 data collection started between 1991 and 1995 when the first questionnaires were sent to 16 years old twins (N = 5778; 90%). The follow-ups were then continued in four different study waves: when twins were 17 (N = 5445; 95%), 18.5 (N = 5427; 94%), 23-27 (N = 5240; 85%) and about 35 years old (N = 4407; 72%). (Kaidesoja et al., 2019.) In this study, four of the study waves from age 16, 18, 23-27 and 35 were used. Those who participated at age 35 were included in the present study.

FinnTwin16 was approved by the Ethical Committee of the Department of Public Health, University of Helsinki, Helsinki University Central Hospital ethical committee, and by the Institutional Review Board of Indiana University.

### 2.2 Parenthood status and parenthood years

In TAM, parenthood status was defined from two questions in each study wave: “Do you have children” and “What year were your children born?”. The detailed question about the children’s birth years was not included in the study wave at age 52 years. Questionnaires at age 22, 32, and 42 were used to obtain the age of becoming a parent. The parenthood year was calculated for each wave from the age of having the first child (birth year of the first child - respondents birth year). For example, if the first child was born in 1997, the parenthood year in 1983 was −14 years, 1989 was −8 years, 1999 +2 years, 2009 +12 years and in 2019 +22 years. Parenthood years were then categorized to 11 categories: −17 or more years before the transition, −16 to −12 years, −11 to - 7 years, −6 to −2 years, −1 to 1 years, 2 to 6 years, 7 to 11 years, 12 to 16 years, 17 to 21 years, 22 to 26 years, and 27 or more years after the transition to parenthood.

For FinnTwin16, parenthood status was defined from the questions about biological children and the children’s birth years at age 35. Based on the children’s birth years, the age of having the first child was calculated. Similar to TAM, parenthood years were then calculated for each wave to and from the age of having the first child. For FinnTwin16, parenthood years were categorized into 8 categories: −17 or more years before the transition to parenthood, −16 to −12 years, −11 to −7 years, −6 to −2 years, −1 to 1 years, 2 to 6 years, 7 to 11 years, and 12 years or more after the transition to parenthood.

In both datasets, respondents who answered “no children” and did not report any children’s birth years in any study wave were categorized as non-parents.

### 2.3 Outcomes

*The frequent alcohol use (FAU)* was assessed with the question “How often do you drink alcohol?” in each study wave across both cohorts. Based on responses to this question, FAU was categorized into two groups: frequent alcohol use and infrequent/no alcohol use. Frequent alcohol use included individuals who reported drinking alcohol at least twice a week. An exception was made for the TAM cohort at age 16, where the options differed from those at other age stages. At age 16, “at least once a week” was the most frequent drinking category. Therefore, at age 16, frequent alcohol use included those who reported drinking alcohol at least once a week.

*The heavy episodic drinking (HED)* was assessed with questions about the frequency of intoxication (TAM 16/22, FT16 16/18) and having five/six or more drinks in a row (TAM: six or more at ages 32/42/52; FT16: more than 5 drinks at ages 25/35). HED was categorized into two classes: frequent HED and infrequent/no HED. In the TAM cohort, the frequent category at age 16 included those who reported being drunk at least four times during the school term. At age 22, the frequent category included those who reported heavy drunkenness at least once a month. At ages 32, 42 and 52, HED was based on the third question in the Alcohol Use Disorders Identification Test (AUDIT), including those who drank six or more drinks in a row once a month or more often. In FinnTwin16, the frequent category included those who reported being drunk at least once a month at ages 16 and 18. At ages 25 and 35, HED was based on the question “At present, how often do you within one occasion drink more than five bottles of beer, or more than a bottle of wine, or more than half a bottle of hard liquor (or a corresponding amount of alcohol)?”. Those who reported drinking more than five drinks at least once a month were categorized as frequent HED.

### 2.4 Covariates

We used the age of becoming a parent, being in a romantic relationship (marriage, cohabitation or dating; yes/no), completed high school (yes/no) and whether the respondent had more than one child (yes/no) from each study wave as covariates.

The age of becoming a parent was calculated from the year of birth of the first child and the respondent’s own year of birth. The age of becoming a parent was also used as a covariate in the analysis comparing parents and nonparents. Because nonparents do not have children and therefore the age of becoming a parent, we calculated the mean age of becoming a parent for nonparents based on the data (TAM: 29 years, FinnTwin16: 28 years).

Being in a romantic relationship, having completed high school, and whether the respondent had more than one child were measured at each study wave. Different measures were used at different ages to determine romantic relationships. In TAM, being in a romantic relationship at age 16 was based on the question “Are you currently dating?”. At age 22, 32, 42 and 52, relationship status was determined by marital status (unmarried, cohabiting, married, divorced/separated, and widowed). In addition, those who were unmarried, divorced/separated, or widowed, were asked whether they were dating (yes/no). Respondents who reported dating, were categorized as being in a relationship. In FinnTwin16, being in a romantic relationship was assessed by dating status at age 18 and the duration of the current relationship at ages 25 and 35. In FinnTwin16, no questions about the relationship or dating were asked at age 16, thus it was inferred from the dating question at age 18.

In TAM, high school completion was asked in each study wave. In FinnTwin16, completed high school was based on the questionnaire about the education at age 35. Those who reported completed high school (or higher education) were coded as having completed high school. Since participants had not yet graduated from high school at the age of 16, their education at age 16 was coded as ‘no basic education’. Having two or more children was also measured dichotomously at each study wave. At age 16, no question on children was asked, but since none of the participants had two or more children based on birth year information, this variable was coded as 0 for all participants.

### 2.5 Statistical analysis

All analyses were performed using IBM SPSS Statistic version 27. Each analysis was performed separately for men and women. We reported frequencies and percentages for categorical variables and means and standard deviations for continuous variables.

We used generalized linear mixed model to examine the association between parenthood years and alcohol use before and after the transition to parenthood. Separate analyses were conducted for the frequent alcohol use and for the heavy episodic drinking. For both outcomes, an unadjusted model was estimated first with parenthood years as the only predictor in the model. For the adjusted model, the age of becoming a parent, romantic relationship, basic education and whether respondent had two or more children were included as covariates. For FinnTwin16, clustering within families (i.e., twin pairs) was taken into account in all analyses. The parenthood year category −1 to 1 years before/after the transition was used as a reference category.

To enhance interpretation of the before vs. after analysis, we additionally performed within subject change analyses using linear mixed models. These compared parenthood year categories −6 to −2 years and +2 to +6 years for both alcohol use outcomes (dichotomized). In the FinnTwin16, a family ID was taking account for the non-independence of observations within families.

To examine the differences between parents’ and non-parents’ alcohol use, we used logistic regression and generalized linear mixed model analysis. We examined these differences (frequent alcohol use and heavy episodic drinking) in five (TAM) and four (FT16) time points from adolescence to mid-adulthood. Analyses were first conducted without any adjustments, and in adjusted model, we included age of becoming a parent (using the sample’s mean age of becoming a parent for non-parents), romantic relationship, basic education and whether respondent had two or more children.

## 3 Results

Descriptive statistics of the study variable are presented in Table 1. In both cohorts, the majority had become parents by their mid-thirties. FAU increased by age in both cohorts among women and men. In TAM, HED peaked at age 16 among women and at age 32 in men, in FinnTwin16 the highest frequency of HED was at age 25 among both women and men.

**Table 1:**
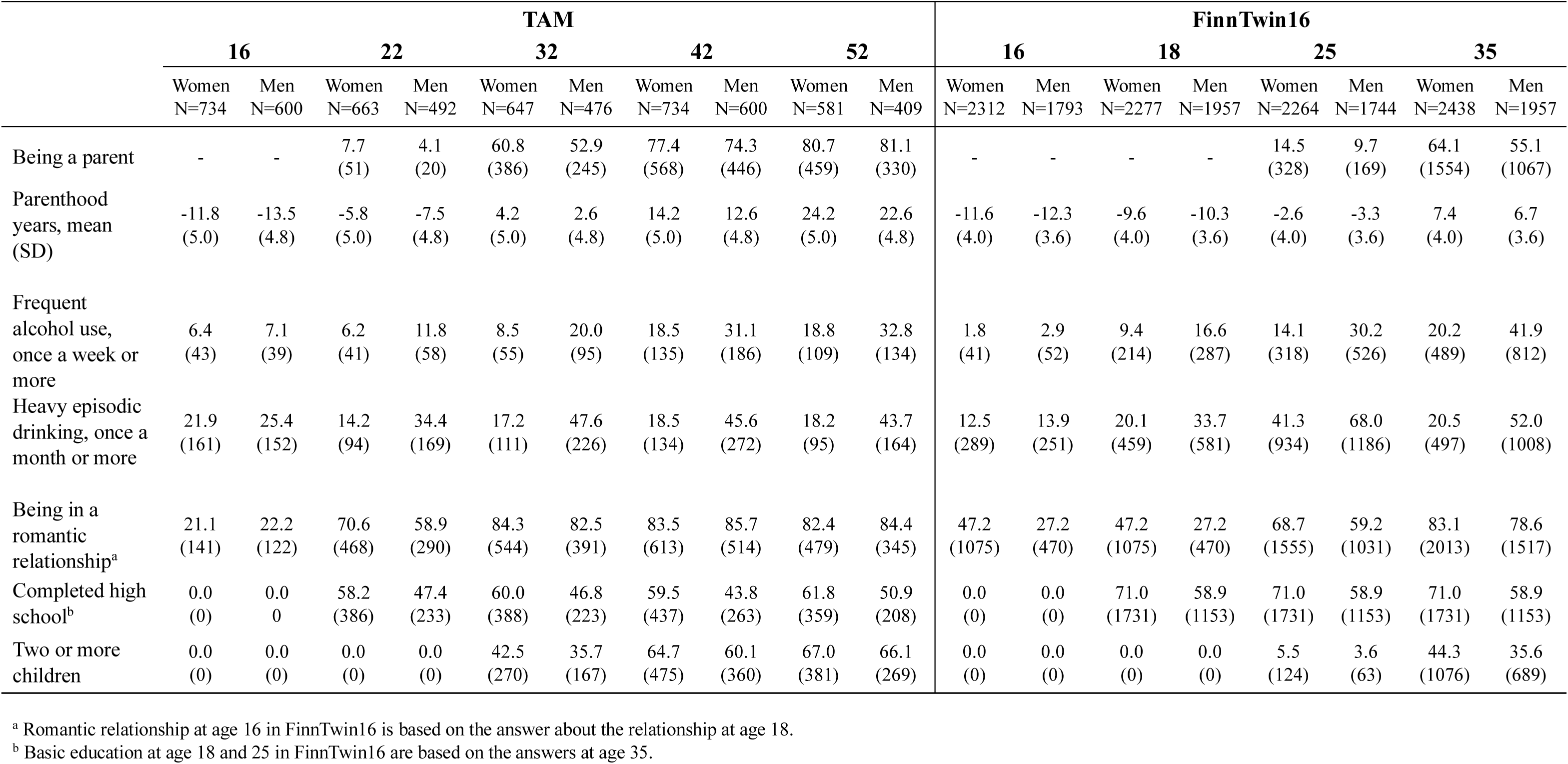
Descriptive statistics.

### 3.1 FAU before and after the transition to parenthood

The generalized linear mixed analysis examined FAU across different parenthood year categories in two Finnish cohorts, using the period from −1 to 1 year around the transition to parenthood as the reference category (Table 2, Figure 1). Among women in the TAM-cohort, no differences were found in frequent alcohol use before the transition to parenthood compared to the reference period +/-1 years around the transition to parenthood in the adjusted model. In FinnTwin16, results showed significantly less frequent alcohol use 12 years or earlier before parenthood compared to the transition period among women. After the transition to parenthood, the results showed an increasing pattern in frequent alcohol use in both cohorts starting from 12 years post-transition in TAM, and 7 years post-transition in FinnTwin16 compared to the immediate transition period in the adjusted model.

**Figure 1.**
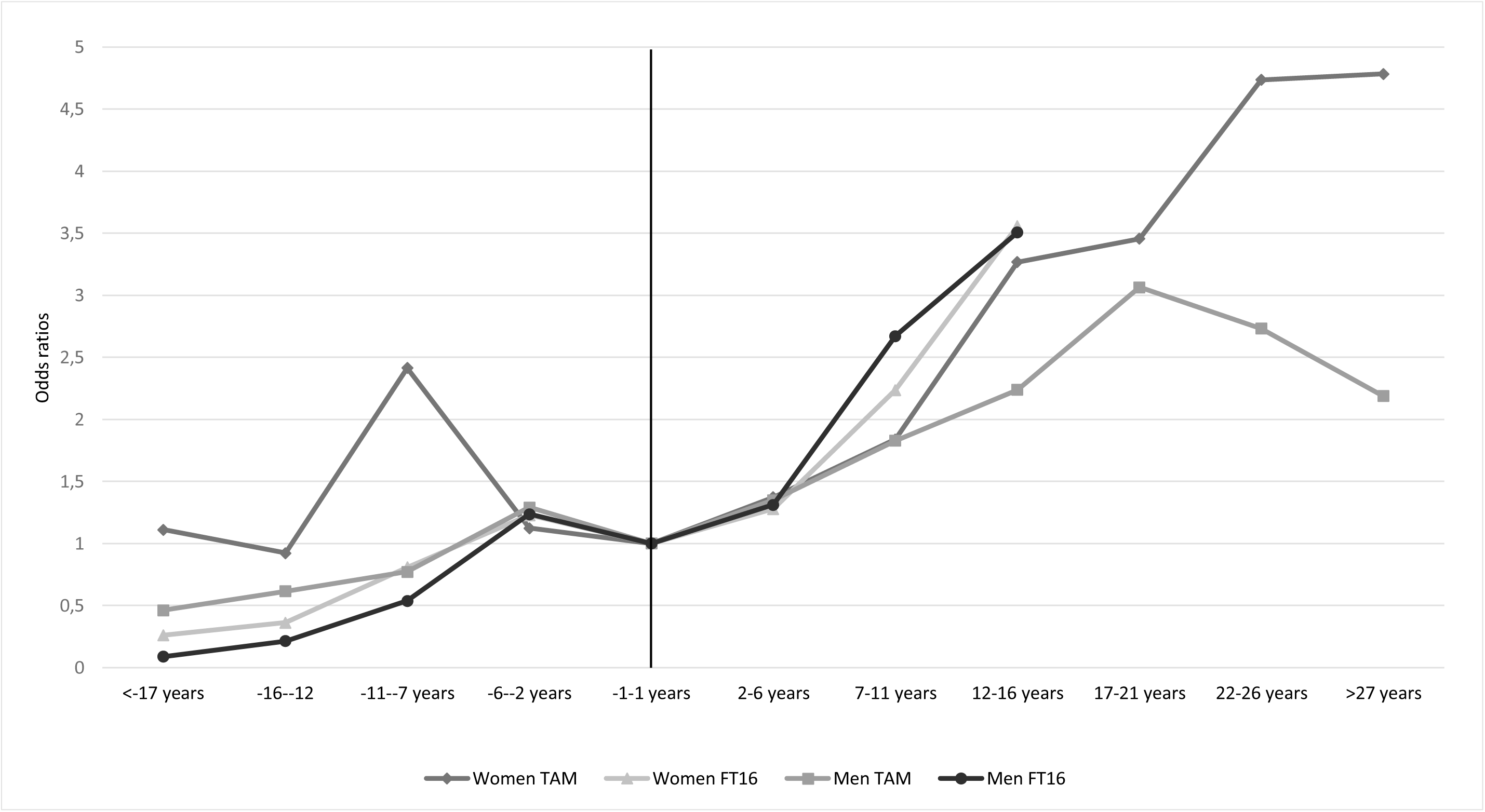
Frequent alcohol use (FAU) before and after becoming a parent in the adjusted model. Odds ratios from generalized linear mixed models with −1 to 1 years before/after parenthood as the reference category.

**Figure 2.**
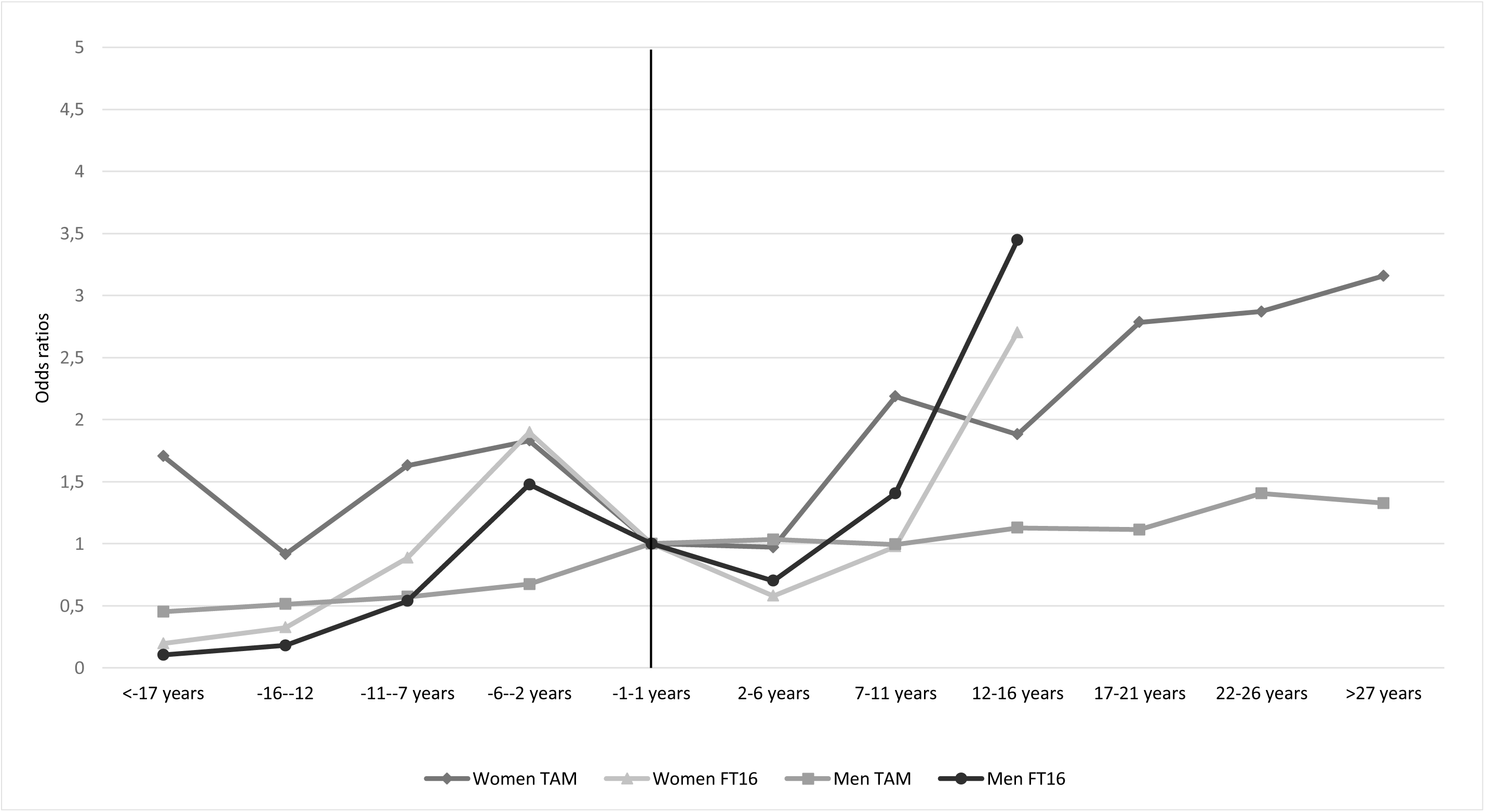
Heavy episodic drinking (HED) before and after becoming a parent in the adjusted model. Odds ratios from generalized linear mixed models with −1 to 1 years before/after parenthood as the reference category.

**Table 2.**
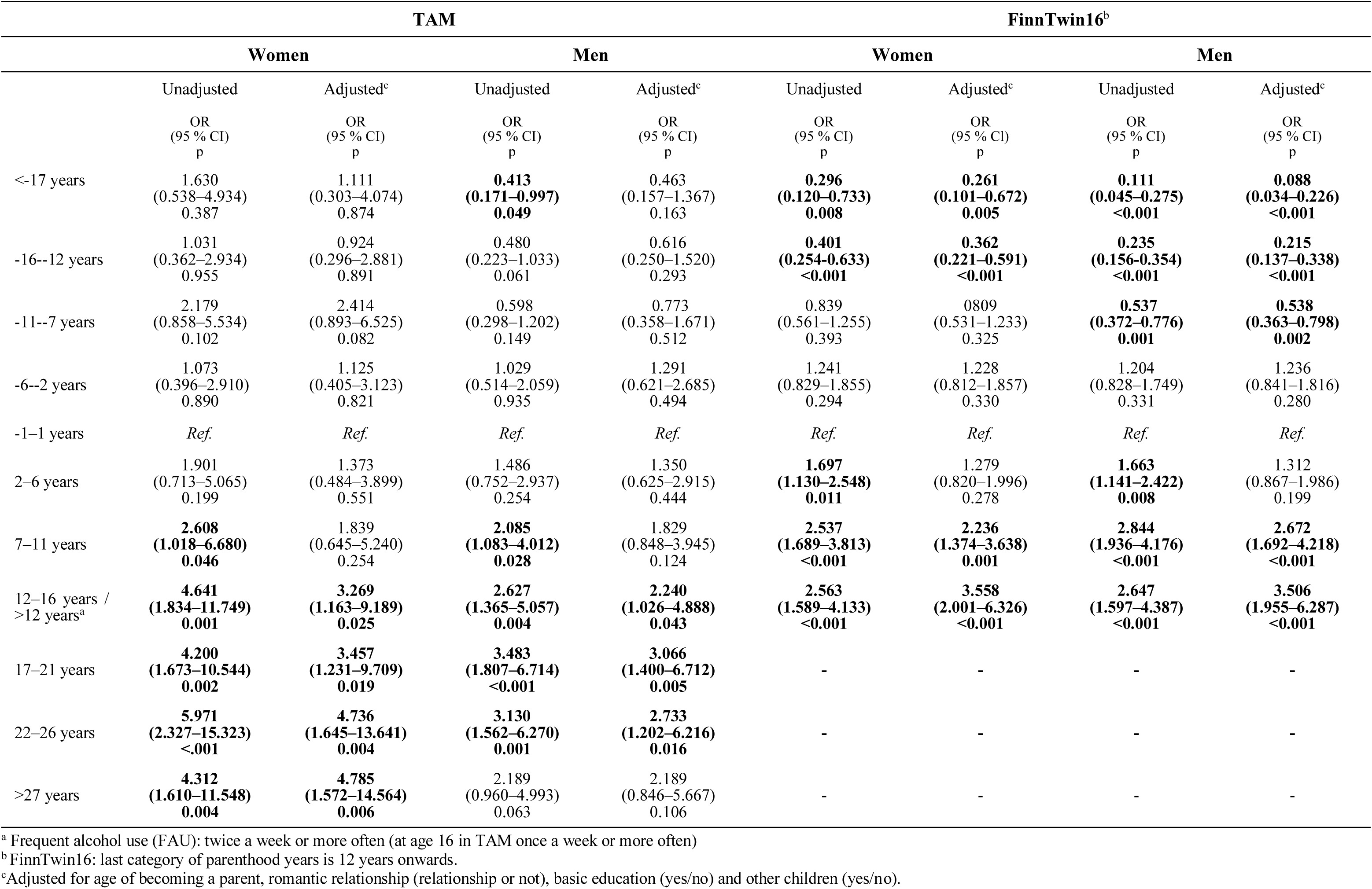
Frequent alcohol use (FAU)^a^ before and after becoming a parent. Odds ratios from generalized linear mixed models with −1-1 years before/after parenthood as the reference category. Only with those participated at age of 42 (TAM) and 35 (FinnTwin16).

In TAM cohort, none of the periods before parenthood showed significant differences in FAU. In FinnTwin16, results showed that fathers drank alcohol significantly less frequently 7 or more years before the transition to parenthood compared to the time of becoming a parent in the adjusted model. After the transition to parenthood, the results showed significant increase in FAU years after the transition in TAM-cohort, particularly from 17 to 26 years post-transition in the adjusted model. In FinnTwin16, fathers drank more frequently after the transition, although in the adjusted model, no differences were found when comparing the time during the transition and immediate years after the transition to parenthood (2 to 6 years after).

### 3.2 HED before and after the transition to parenthood

Among women in TAM cohort, the results showed no significant differences in the odds of HED before the transition to parenthood compared to the reference period (Table 3). In FinnTwin16, HED increased as individuals approached parenthood; the odds of HED were significantly lower until 12 years before parenthood and significantly higher 6 to 2 years before the transition in the adjusted model. After the transition to parenthood, the odds of HED were significantly higher 7 to 11 years post-transition, and from 17 years onwards in TAM. In FinnTwin16, the results in the adjusted model showed that immediately after the transition (2 to 6 years after the transition, OR=0.579, p<0.003), mother’s HED was significantly lower than during the transition period. However, the odds of HED were significantly higher when the first child was born 12 years ago or earlier (OR=2.703, p<0.001).

**Table 3:** Heavy episodic drinking (HED)^a^ before and after becoming a parent. Odds ratios from generalized linear mixed models with −1-1 years before/after parenthood as the reference category. Only with those participated at age of 42 (TAM) and 35 (FinnTwin16).

|  | TAM |  |  |  | FinnTwin16<sup>b</sup> |  |  |  |
| --- | --- | --- | --- | --- | --- | --- | --- | --- |
|  | WOMEN |  | MEN |  | WOMEN |  | MEN |  |
|  | Unadjusted | Adjusted<sup>c</sup> | Unadjusted | Adjusted<sup>c</sup> | Unadjusted | Adjusted<sup>c</sup> | Unadjusted | Adjusted<sup>c</sup> |
|  | OR<br>(95 % CI)<br>p | OR<br>(95 % CI)<br>p | OR<br>(95 % CI)<br>p | OR<br>(95 % CI)<br>p | OR<br>(95 % CI)<br>p | OR<br>(95 % CI)<br>p | OR<br>(95 % CI)<br>p | OR<br>(95 % CI)<br>p |
| <-17 years | **2.496**<br>**(1.202–5.184)**<br>**0.014** | 1.705<br>(0.668–4.351)<br>0.264 | **0.367**<br>**(0.193–0.699)**<br>**0.002** | 0.452<br>(0.200–1.020)<br>0.056 | **0.316**<br>**(0.175–0.572)**<br>**<0.001** | **0.196**<br>**(0.105–0.369)**<br>**<0.001** | **0.131**<br>**(0.074–0.233)**<br>**<0.001** | **0.104**<br>**(0.056–0.196)**<br>**<0.001** |
| -16--12 years | 1.326<br>(0.674–2.608)<br>0.414 | 0.916<br>(0.415–2.023)<br>0.829 | **0.415**<br>**(0.235–0.735)**<br>**0.003** | 0.512<br>(0.259–1.015)<br>0.055 | **0.461**<br>**(0.335–0.633)**<br>**<0.001** | **0.325**<br>**(0.229–0.461)**<br>**<0.001** | **0.202**<br>**(0.143–0.285)**<br>**<0.001** | **0.180**<br>**(0.122–0.265)**<br>**<0.001** |
| -11--7 years | **2.063**<br>**(1.113–3.825)**<br>**0.022** | 1.629<br>(0.815–3.256)<br>0.168 | **0.555**<br>**(0.324–0.950)**<br>**0.032** | 0.571<br>(0.318–1.026)<br>0.061 | 1.047<br>(0.785–1.395)<br>0.756 | 0.886<br>(0.657–1.196)<br>0.428 | **0.539**<br>**(0.391–0.743)**<br>**<0.001** | **0.537**<br>**(0.381–0.756)**<br>**<0.001** |
| -6--2 years | **1.979**<br>**(1.061–3.691)**<br>**0.032** | 1.830<br>(0.964–3.475)<br>0.065 | 0.663<br>(0.375–1.171)<br>0.157 | 0.674<br>(0.375–1.213)<br>0.188 | **2.014**<br>**(1.502–2.699)**<br>**<0.001** | **1.898**<br>**(1.409–2.557)**<br>**<0.001** | **1.430**<br>**(1.017–2.009)**<br>**0.040** | **1.477**<br>**(1.042–2.094)**<br>**0.028** |
| -1–1 years | *Ref.* | *Ref.* | *Ref.* | *Ref.* | *Ref.* | *Ref.* | *Ref.* | *Ref.* |
| 2–6 years | 0.689<br>(0.335–1.416)<br>0.311 | 0.970<br>(0.458–2.055)<br>0.937 | 0.938<br>(0.528–1.667)<br>0.828 | 1.033<br>(0.547–1.953)<br>0.919 | **0.499**<br>**(0.358–0.696)**<br>**<0.001** | **0.579**<br>**(0.405–0.829)**<br>**0.003** | **0.645**<br>**(0.456–0.912)**<br>**0.013** | 0.702<br>(0.479–1.030)<br>0.071 |
| 7–11 years | 1.424<br>(0.744–2.723)<br>0.285 | **2.186**<br>**(1.041–4.588)**<br>**0.039** | 0.904<br>(0.519–1.575)<br>0.721 | 0.992<br>(0.519–1.898)<br>0.981 | **0.634**<br>**(0.450–0.892)**<br>**0.009** | 0.977<br>(0.644–1.482)<br>0.913 | 0.937<br>(0.655–1.340)<br>0.722 | 1.406<br>(0.914–2.163)<br>0.121 |
| 12–16 years /<br>>12 years<sup>a</sup> | 1.118<br>(0.565–2.213)<br>0.749 | 1.880<br>(0.869–4.070)<br>0.109 | 1.014<br>(0.576–1.784)<br>0.962 | 1.128<br>(0.575–2.212)<br>0.727 | 1.341<br>(0.905–1.989)<br>0.144 | **2.703**<br>**(1.672–4.369)**<br>**<0.001** | **1.672**<br>**(1.020–2.740)**<br>**0.041** | **3.445**<br>**(1.954–6.072)**<br>**<0.001** |
| 17–21 years | 1.742<br>(0.916–3.313)<br>0.090 | **2.785**<br>**(1.324–5.856)**<br>**0.007** | 1.024<br>(0.574–1.824)<br>0.937 | 1.113<br>(0.562–2.202)<br>0.759 | - | - | - | - |
| 22–26 years | 1.679<br>(0.839–3.359)<br>0.143 | **2.871**<br>**(1.300–6.339)**<br>**0.009** | 1.209<br>(0.647–2.260)<br>0.551 | 1.405<br>(0.675–2.927)<br>0.363 | - | - | - | - |
| >27 years | 1.945<br>(0.952–3.974)<br>0.068 | **3.158**<br>**(1.397–7.138)**<br>**0.006** | 1.178<br>(0.549–2.531)<br>0.674 | 1.326<br>(0.558–3.150)<br>0.523 | - | - | - | - |

<sup>a</sup> Heavy episodic drinking (HED): once a month or more often
<sup>b</sup> FinnTwin16: last category of parenthood years is 12 years onwards.
<sup>c</sup> Adjusted model: adjusted for age of becoming a parent, relationship status (dichotomous, relationship or not), basic education (yes/no) and other children (yes/no).

Among men, similar patterns of HED were found in both cohorts years before becoming parents. In TAM, fathers’ HED increased to the transition to parenthood, but these results disappeared after the adjustments. In FinnTwin16, an increasing pattern of HED was found, with the odds of HED significantly lower 7 or more years before parenthood but significantly higher in the 6 to 2 years immediately before the transition in adjusted model. After the transition to parenthood, no significant associations were found among men in TAM cohort. However, in FinnTwin16, the HED was significantly higher twelve years after the transition to parenthood than during the transition.

### 3.3 Within person changes

Comparing the period 2–6 years before becoming a parent to the period 2–6 years after in a within-individual repeated-measurements analysis no significant changes in FAU were observed in either cohort among either women or men (Table 4). Regarding HED, there was a significant reduction among women in both cohorts and among men in FinnTwin16 cohort, while no significant within-person changes were found in men in TAM.

**Table 4.**
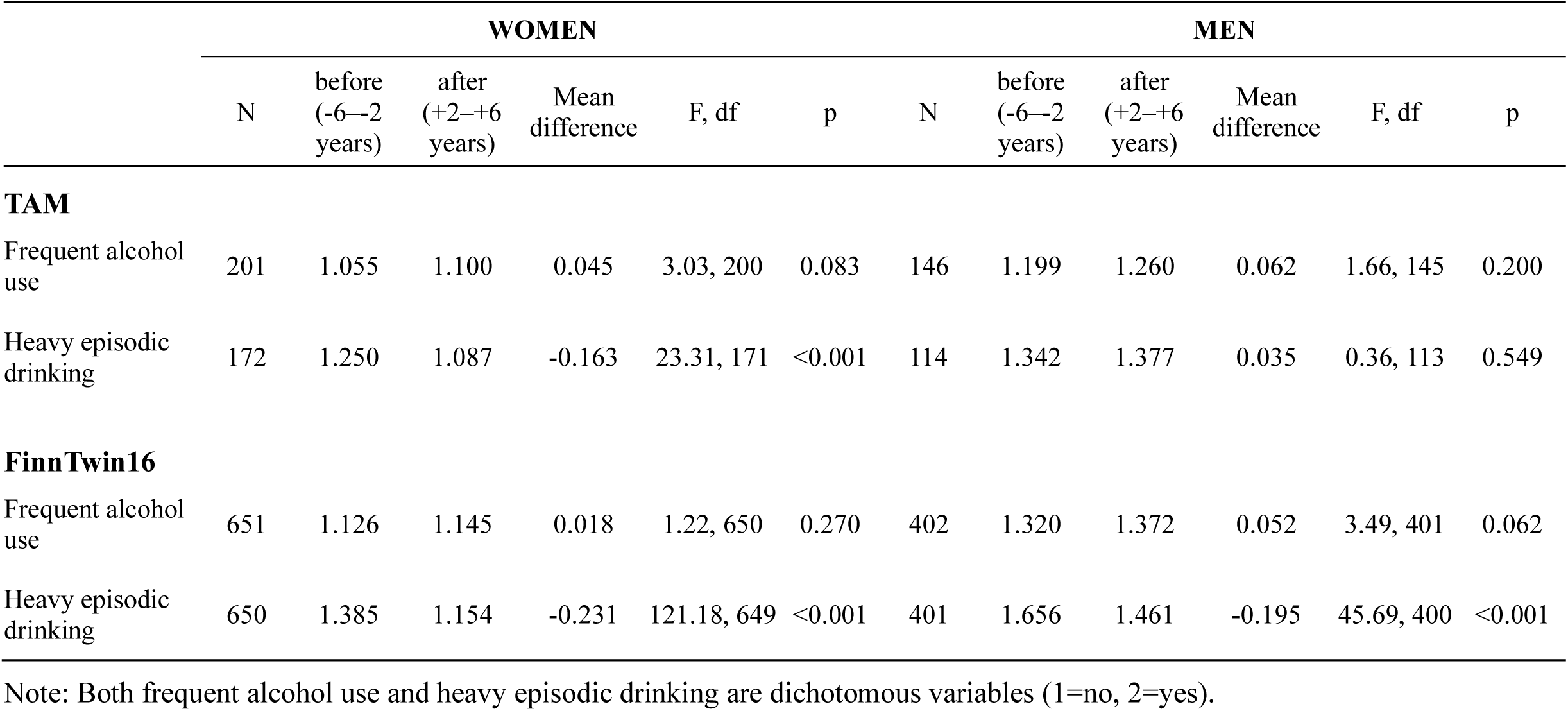
Within subject changes of frequent alcohol use and heavy episodic drinking from before (–6 to –2 years) to after (+2 to +6 years) becoming a parent. Estimated marginal means and mean differences from linear mixed models.

|  | WOMEN |  |  |  |  |  | MEN |  |  |  |  |  |
| --- | --- | --- | --- | --- | --- | --- | --- | --- | --- | --- | --- | --- |
|  | N | before<br>(−6−2<br>years) | after<br>(+2−+6<br>years) | Mean<br>difference | F, df | p | N | before<br>(−6−2<br>years) | after<br>(+2−+6<br>years) | Mean<br>difference | F, df | p |
| **TAM** |  |  |  |  |  |  |  |  |  |  |  |  |
| Frequent alcohol use | 201 | 1.055 | 1.100 | 0.045 | 3.03, 200 | 0.083 | 146 | 1.199 | 1.260 | 0.062 | 1.66, 145 | 0.200 |
| Heavy episodic drinking | 172 | 1.250 | 1.087 | −0.163 | 23.31, 171 | <0.001 | 114 | 1.342 | 1.377 | 0.035 | 0.36, 113 | 0.549 |
| **FinnTwin16** |  |  |  |  |  |  |  |  |  |  |  |  |
| Frequent alcohol use | 651 | 1.126 | 1.145 | 0.018 | 1.22, 650 | 0.270 | 402 | 1.320 | 1.372 | 0.052 | 3.49, 401 | 0.062 |
| Heavy episodic drinking | 650 | 1.385 | 1.154 | −0.231 | 121.18, 649 | <0.001 | 401 | 1.656 | 1.461 | −0.195 | 45.69, 400 | <0.001 |

Note: Both frequent alcohol use and heavy episodic drinking are dichotomous variables (1=no, 2=yes).

### 3.4 Differences between parents and nonparents

Logistic regression analysis in TAM and generalized linear mixed model analysis in FinnTwin16 was conducted to examine the differences of alcohol use between parents and nonparents at five (TAM) or four (FT16) time points (Table 5 & 6). In TAM, mothers were significantly less likely to engage in FAU than nonmothers at age 32 and 42. After the adjustments, only difference persisted at age 42, with mothers drinking less frequently than nonmothers. In FinnTwin16, the odds of FAU were significantly lower for mothers compared to nonmothers at age 25, but only in the unadjusted model. Among men in TAM cohort, no significant differences were found in the frequency of alcohol use between parents and nonparents. In FinnTwin16, fathers reported more frequent drinking than nonfathers at age 25 in adjusted model.

**Table 5.**
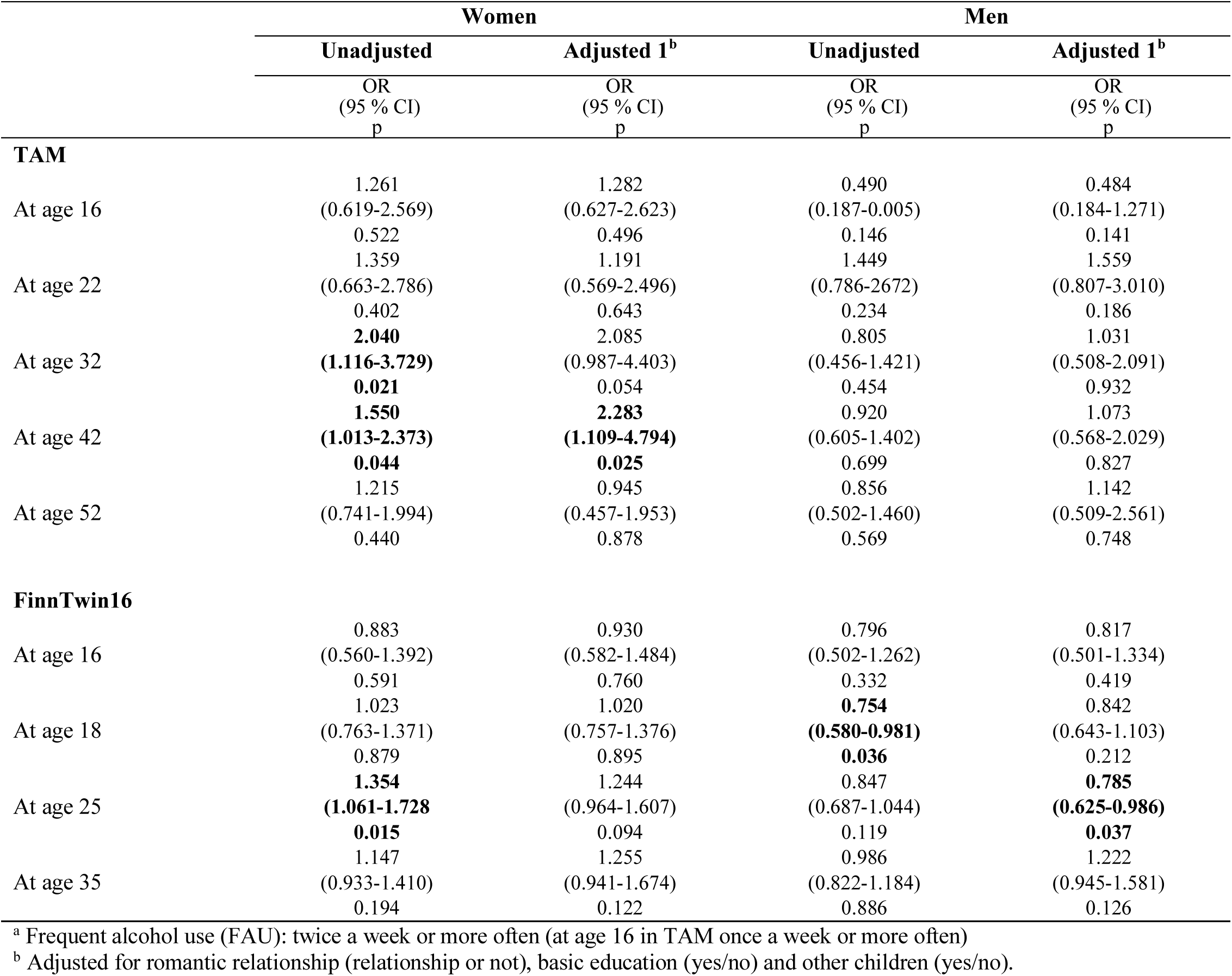
Odds ratios for frequent alcohol use^a^ comparing nonparents to parents (reference group) across different timepoints. Results from logistic regression models (TAM) and generalized linear mixed models (FinnTwin16).

**Table 6.**
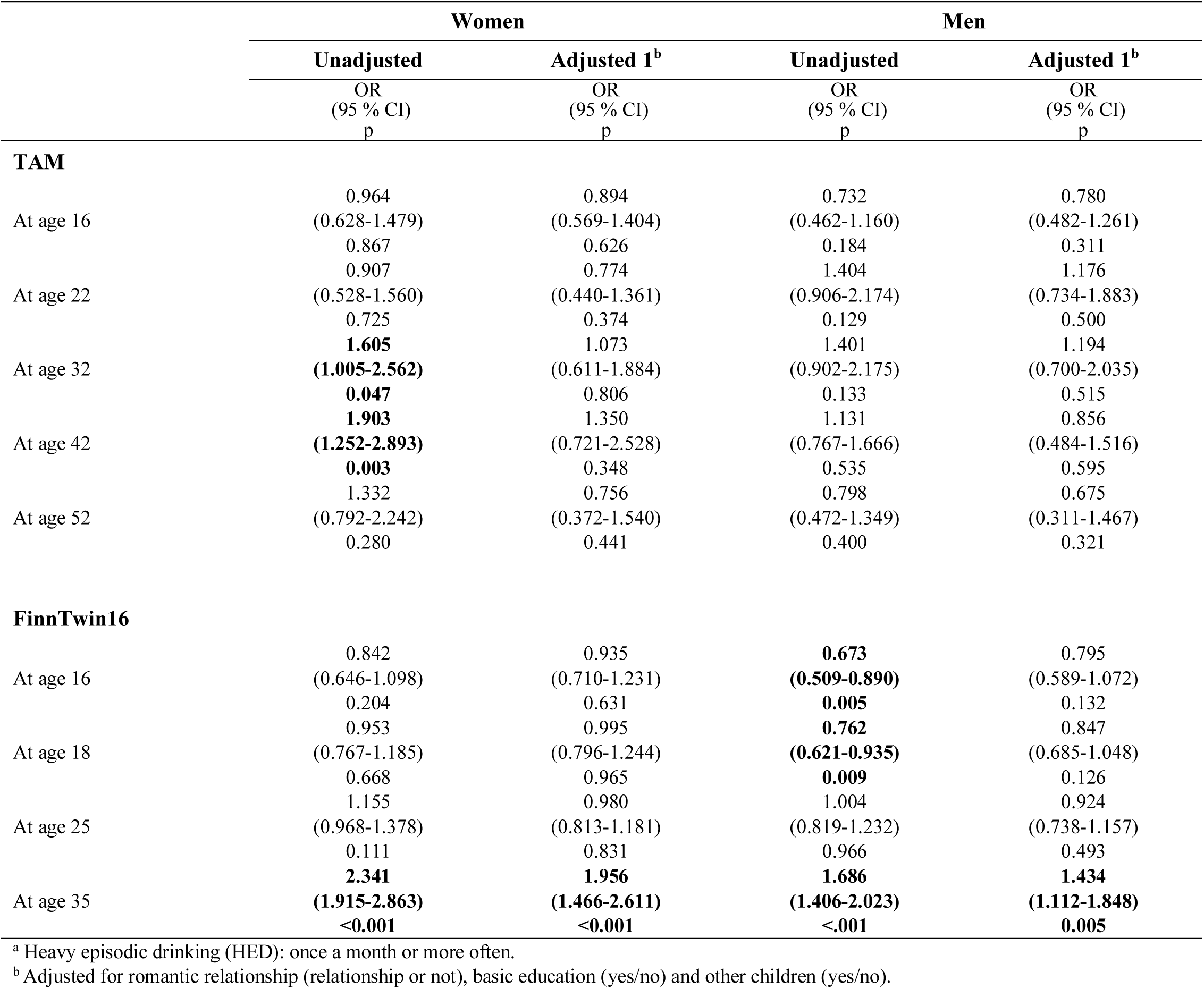
Odds ratios for heavy episodic drinking^a^ comparing nonparents to parents (reference group) across different timepoints. Results from logistic regression models (TAM) and generalized linear mixed models (FinnTwin16).

|  | Women |  | Men |  |
| --- | --- | --- | --- | --- |
|  | Unadjusted | Adjusted 1<sup>b</sup> | Unadjusted | Adjusted 1<sup>b</sup> |
|  | OR<br>(95 % CI)<br>p | OR<br>(95 % CI)<br>p | OR<br>(95 % CI)<br>p | OR<br>(95 % CI)<br>p |
| **TAM** |  |  |  |  |
| At age 16 | 0.964 | 0.894 | 0.732 | 0.780 |
|  | (0.628-1.479) | (0.569-1.404) | (0.462-1.160) | (0.482-1.261) |
|  | 0.867 | 0.626 | 0.184 | 0.311 |
| At age 22 | 0.907 | 0.774 | 1.404 | 1.176 |
|  | (0.528-1.560) | (0.440-1.361) | (0.906-2.174) | (0.734-1.883) |
|  | 0.725 | 0.374 | 0.129 | 0.500 |
| At age 32 | **1.605** | 1.073 | 1.401 | 1.194 |
|  | **(1.005-2.562)** | (0.611-1.884) | (0.902-2.175) | (0.700-2.035) |
|  | **0.047** | 0.806 | 0.133 | 0.515 |
| At age 42 | **1.903** | 1.350 | 1.131 | 0.856 |
|  | **(1.252-2.893)** | (0.721-2.528) | (0.767-1.666) | (0.484-1.516) |
|  | **0.003** | 0.348 | 0.535 | 0.595 |
| At age 52 | 1.332 | 0.756 | 0.798 | 0.675 |
|  | (0.792-2.242) | (0.372-1.540) | (0.472-1.349) | (0.311-1.467) |
|  | 0.280 | 0.441 | 0.400 | 0.321 |
| **FinnTwin16** |  |  |  |  |
| At age 16 | 0.842 | 0.935 | **0.673** | 0.795 |
|  | (0.646-1.098) | (0.710-1.231) | **(0.509-0.890)** | (0.589-1.072) |
|  | 0.204 | 0.631 | **0.005** | 0.132 |
| At age 18 | 0.953 | 0.995 | **0.762** | 0.847 |
|  | (0.767-1.185) | (0.796-1.244) | **(0.621-0.935)** | (0.685-1.048) |
|  | 0.668 | 0.965 | **0.009** | 0.126 |
| At age 25 | 1.155 | 0.980 | 1.004 | 0.924 |
|  | (0.968-1.378) | (0.813-1.181) | (0.819-1.232) | (0.738-1.157) |
|  | 0.111 | 0.831 | 0.966 | 0.493 |
| At age 35 | **2.341** | **1.956** | **1.686** | **1.434** |
|  | **(1.915-2.863)** | **(1.466-2.611)** | **(1.406-2.023)** | **(1.112-1.848)** |
|  | **<0.001** | **<0.001** | **<.001** | **0.005** |

<sup>a</sup> Heavy episodic drinking (HED): once a month or more often.
<sup>b</sup> Adjusted for romantic relationship (relationship or not), basic education (yes/no) and other children (yes/no).

Regarding HED, mothers were less likely to engage in heavy episodic drinking than nonmothers at age 32 and

42 in TAM cohort. However, after adjusting for the model, these significant results disappeared. In FinnTwin16, a similar finding was found among mothers at age 35, with this result remaining significant in the adjusted model. In TAM, no differences were found among fathers and nonfathers. In FinnTwin16, fathers exhibited higher rates of HED compared to nonfathers at ages 16 and 18, but lower rates at age 35, while the result regarding age 35 remained significant after the adjustments.

## 4 Discussion

The present study examined frequent alcohol use and heavy episodic drinking in relation to parenthood across two different Finnish cohorts, spanning from adolescence to mid-adulthood, and revealed distinct change patterns based on gender, cohort and type of alcohol use. Across both cohorts FAU remained relatively stable around the transition period, while increased later on during the life course. With the exception of men in the TAM cohort, HED declined from before to after the transition period, but increased beyond those levels in the later years of parenthood. In the FinnTwin16 cohort, nonparents reported HED more often than parents at age 35. Otherwise, there were only a few differences between parents and nonparents, suggesting that parenthood may not strongly differentiate alcohol use patterns.

Before the transition to parenthood, an increasing pattern with alcohol use was found, particularly in the FinnTwin16 cohort. Among both women and men in FinnTwin16, FAU and HED were significantly lower years before the transition to parenthood and rose steadily as the transition approached. HED was already significantly more likely during the 6 to 2 years before parenthood than during the transition. These findings aligns with previous research showing that alcohol use typically increases through adolescence and early adulthood (Maggs & Schulenberg, 2004). Previous studies have also showed that alcohol consumption tends to peak right before major life transitions that bring increased responsibilities (Arria et al., 2016; Staff et al., 2010). In the TAM cohort, fewer differences were observed as only among men, HED was more likely in some pre-parenthood periods.

Our study reveals nuanced patterns of alcohol use following the transition to parenthood. Among women and men, FAU remained relatively stable during the early post-transition period (2 to 6 years after becoming a parent compared to the transition period), consistent with some previous studies (Matusiewicz et al., 2016). However, HED declined during this period among women in both cohorts and among men in FinnTwin16, indicating a short-term reduction in episodic drinking behavior following the transition to parenthood. Especially these changes were evident in the within subject analyses comparing to the period of 6 to 2 years before the transition. These findings suggest that while occasional alcohol use may continue, parents tend to moderate heavier drinking behaviors in the early years of parenting, potentially reflecting increased responsibilities or changes in social behavior.

Beyond this early parenthood phase, longer term results showed that alcohol use became more frequent and HED more likely over time in both cohorts. This trend may reflect growing opportunities for socializing and alcohol consumptions as children grow older and parenting demands lessen. However, it may also partly reflect general age-related trends and cultural changes in alcohol consumption (Britton et al., 2015; Tigerstedt et al., 2020). Overall, our findings indicate that the transition to parenthood is associated with short-term reductions in heavy episodic drinking, particularly among women, but that these effects are time limited. In the longer term, both mothers and fathers showed increasing alcohol use, highlighting the importance of considering both immediate and extended time frames when assessing the relationship between parenthood and alcohol use.

We also examined the differences between parents and nonparents. In TAM cohort, mothers reported significantly less frequent alcohol use than non-mothers at age 42, while in FinnTwin16, mothers were less likely to engage in HED at age 35. These observed differences in alcohol use between mother and non-mothers may reflect the influence of the prime child-rearing years and early parenthood. Becoming a parent often introduces increased caregiving responsibilities and lifestyle changes, which may lead mothers to adopt more cautious drinking behaviors during this period (Paradis et al., 2011). In addition, societal norms and expectation surrounding motherhood may contribute to these differences (Ujhelyi Gomez et al., 2022). For men, only two differences were found in FinnTwin16, were fathers reported more frequent alcohol use at age 25, but were less likely to engage in heavy episodic drinking at age 35. Taken together, the overall differences between parents and non-parents were limited in both women and men. This suggest that parenthood alone may not be a strong predictor of alcohol use during the life course, and that observed differences may be more shaped by the timing, individual trajectories, or broader social and demographic contexts. These findings should also be interpreted considering possible selection effects, as earlier alcohol use may influence associated who becomes a parent in the first place (Rose et al., 2022).

Some differences in alcohol consumption patterns regarding frequent alcohol use and heavy episodic drinking were revealed, suggesting that factors influencing occasional heavy drinking versus regular alcohol consumption may vary. Before parenthood, individuals may moderate their overall frequency of drinking as they anticipate future responsibilities, whereas occasional heavy drinking episodes could still occur in social contexts. After becoming parents, changes in social dynamics, stress levels, and coping mechanisms may lead to differing patterns: more frequent drinking could reflect increased social opportunities or stress relief, while reductions in HED might result from shifts towards more stable routines and fewer opportunities for intense socializing.

### 4.1 Limitations

We acknowledge several study limitations. First, the gaps between follow-up waves mean that we may have missed important immediate changes in behavior, particularly those occurring between the ten-year intervals in the TAM study and in the last interval in the FinnTwin16. These gaps mean that various life events and changes occurring between the waves may have influenced the outcomes, thus potentially introducing confounding that could not be accounted for. Additionally, the original datasets did not focus specifically on the transition to parenthood, limiting the causal inferences that can be drawn from the data.

Second, the difference in the end points of the cohorts introduces a limitation: the TAM study follows participants until age 52, while the FinnTwin16 study ends at age 35. This discrepancy means that we have a longer follow-up period for TAM, capturing their experiences through middle adulthood, while the FinnTwin16 participants are only followed through early adulthood. Many parents in the FinnTwin16 cohort may still be in the toddlerhood phase of parenthood, whereas the TAM participants at age 52 are in a different life stage with potentially older or adult children. As a result, direct comparisons between the cohorts were not possible beyond early adulthood.

There are also couple of limitations regarding the measurements. The use of self-reported questionnaires introduces potential biases. Participants might underreport or overreport their alcohol consumption, which could affect the accuracy of our results. We also did not have information whether being childless was voluntary or involuntary, such as due to illness or social circumstances. This distinction is important, as voluntary or involuntary childlessness can have different psychological and social implications, potentially influencing alcohol use patterns differently.

Furthermore, our sample consists of two specific cohorts from Finland, which may limit the generalizability of our findings to other populations or cultural contexts. For example, the Finnish context is shaped not only by the Nordic model’s emphasis on family-friendly policies, such as parental leave, affordable childcare, and gender equality, but also by the Finnish drinking culture. Patterns of alcohol use in Finland have historically been characterized by heavy episodic drinking, although drinking practices have shifted over the years (Tigerstedt et al., 2020). These cultural and policy contexts may influence parents’ alcohol use differently compared to countries with less supportive family policies or different drinking norms.

## 5 Conclusion

Using two longitudinal cohort studies from adolescence to adulthood/mid-adulthood, we examined the long-term changes in alcohol use before and after the transition to parenthood. Among those who became parents, we found an increase in alcohol use years before becoming a parent, followed by a short-term reduction – especially in heavy episodic drinking – during the early years after the transition. In the longer term, however, alcohol use increased again, with patterns varying according to gender, cohort, and type of alcohol use. These findings suggest that while parenthood may temporarily suppress risky drinking behaviors, long-term trajectories reflect broader age-related and social patterns rather than parenthood itself.

## Data Availability

The information on the datasets analyzed during the current study is available on the following web pages www.thl.fi/en/tam (TAM cohort) and http://www.helsinki.fi/fi/tutkimusryhmat/kaksostutkimus (Finntwin16).The data are available upon request. Data requests are reviewed at the Finnish Institute for Health and Welfare (THL) (for TAM) and the Institute for Molecular Medicine Finland (FIMM) Data Access Committee (DAC) (for FinnTwin16) for authorized researchers who have IRB/ethics approval and an institutionally approved study plan. For more details, please contact THL (noora.berg{at}thl.fi /olli.kiviruusu{at}thl.fi) and the FIMM DAC (fimm-dac{at}helsinki.f).

